# Detection of cognitive deficits years prior to clinical diagnosis across neurological conditions

**DOI:** 10.1101/2024.07.10.24310213

**Authors:** Xin You Tai, Sofia Toniolo, David Llewellyn, Cornelia M. van Duin, Masud Husain, Sanjay Manohar

## Abstract

**Importance:** Understanding the cognitive trajectory of a neurological disease can provide important insight on underlying mechanisms and disease progression. Cognitive impairment is now well established as beginning many years before the diagnosis of Alzheimer’s disease, but pre-diagnostic profiles are unclear for other neurological conditions that may be associated with cognitive impairment.

**Objective:** Compare pre-diagnostic and post-diagnostic cognition and global brain volume in ischaemic stroke, focal epilepsy, Parkinson’s disease, multiple sclerosis, motor neurone disease (amyotrophic lateral sclerosis) and migraine using time-to-diagnosis and time-from-diagnosis data in relation to time of assessment.

**Design:** Analysis the prospective UK Biobank cohort with study baseline assessment performed between 2006-2010 and participants followed until 2021.

**Setting:** Multicenter, population-based study.

**Participants:** Sample of 497,252 participants, aged between 38 and 72 years, at baseline with an imaging sub-sample of 42,468 participants.

**Exposure:** Participants with each neurological condition were compared to a healthy control group.

**Main outcomes and measures:** A continuous measure of executive function and magnetic resonance imaging brain measures of total grey matter and hippocampal volume.

**Results:** Of the 497,252 participants (226,206 [45.5%] men, mean [SD] age, 57.5[8.1] years), 12,755 had ischaemic stroke, 6,758 had a diagnosis of focal epilepsy, 3,315 had Parkinson’s disease, 2,315 had multiple sclerosis, 559 had motor neurone disease and 18,254 had migraine either at study baseline or diagnosed during the follow-up period. Apart from motor neurone disease, all conditions had lower *pre-diagnosis* executive function compared to controls (assessment performed median 7.4 years before diagnosis). Participants with focal epilepsy and multiple sclerosis showed a gradual worsening in executive function up to 15 years prior to diagnosis, while ischaemic stroke was characterised by a modest decline for a few years followed by a substantial reduction at the time of diagnosis. By contrast, participants with migraine showed improved post-diagnosis cognitive scores. Pre-diagnosis MRI grey matter volume was lower than controls for stroke, Parkinson’s disease and multiple sclerosis (scans performed median 1.7 years before diagnosis), while other conditions had lower volumes post-diagnosis.

**Conclusion:** These cognitive trajectory models reveal disease-specific temporal patterns, including a long cognitive prodrome associated with focal epilepsy and multiple sclerosis. The findings may help to prioritise risk management of individual diseases and inform clinical decision-making.

**Key Points:** *Question:* What is the pre-diagnosis cognitive profile across neurological conditions of ischaemic stroke, focal epilepsy, Parkinson’s disease, multiple sclerosis, motor neurone disease and migraine?

*Findings:* This cohort study of 495 149 participants, aged between 38 and 72 years, identified gradual worsening of cognition up to 15 years prior to clinical diagnosis in participants with focal epilepsy and multiple sclerosis while stroke was associated with a modest decline for a few years followed by a substantial reduction at the time of diagnosis. Migraine was associated with post-diagnosis improvement in cognitive scores.

*Meaning:* Cognitive trajectory models identify disease-specific temporal patterns that may help to prioritise risk management of individual diseases and inform clinical decision-making.

## Introduction

The cognitive profile of a neurological disease across time can provide important insights into underlying mechanisms, response to treatment, disease progression and prognosis. In Alzheimer’s disease, cognitive impairment may occur decades prior to the diagnosis^1^ which reflects ongoing accumulation of neurodegenerative protein and associated cellular dysfunction and death.^2,3^ Following diagnosis, comparison of cognitive trajectories provides an index of response to treatment such as cholinesterase inhibitors^4^ and the recently described amyloid immunotherapies.^5,6^ In other neurological conditions that may cause cognitive impairment, however, pre- and post-diagnosis cognitive profiles are largely unknown.

Longitudinal evaluation is crucial to map cognitive trajectories. Once a diagnosis has been given, individuals with a condition can be followed up with repeat cognitive measurements to understand the post-diagnosis cognitive changes.^7^ Delineating cognition before the disease is diagnosed, however, is less straightforward. The current pre-diagnosis cognitive model of Alzheimer’s disease^1^ has been classically derived from autosomal dominant Alzheimer’s disease, leveraging families with offspring that have 50% chance of developing the disease.^8,9^ This cognitive model is corroborated by longitudinal studies of individuals with mild cognitive impairment who progress to develop Alzheimer’s disease.^10,11^

For other neurological conditions, pre-diagnostic assessment is often not feasible, and even if it is, longitudinal evaluation is resource-intensive and logistically difficult. Therefore, many important questions remain unanswered. For example, what is the potential and timeframe of post-stroke cognitive recovery? Many studies describe post-stroke cognitive impairment^12^ but few report data from multiple time points^13^ or performance after one year.^14^ Do individuals with focal epilepsy have cognitive impairment prior to starting anti-epileptic medications? One group identified worse memory scores present in new cases of focal epilepsy before treatment initiation,^15^ but precisely when these difficulties begin is unclear. Similarly, what is the cognitive impact of multiple sclerosis and does this improve with treatment? Recent evidence suggests that cortical lesions exist in the early pathogenesis of some people with multiple sclerosis which may affect cognitive function.^16^ However, there is little corroboration outside of small studies. One important related consideration is that cognitive decline may be coupled with reduction in brain volume observed in these conditions.^17–19^

In this study, we use a data-driven approach to understand *both* pre- and post-diagnosis cognitive profiles and neuroimaging data from to the UK Biobank prospective cohort across a range of neurological diseases. Specifically, in relation to date of diagnosis, we model the time-course and magnitude of cognitive impairment across individuals with ischaemic stroke, focal epilepsy, Parkinson’s disease, multiple sclerosis, motor neurone disease (amyotrophic lateral sclerosis) and migraine, compared to healthy controls. We hypothesized that conditions such as focal epilepsy and Parkinson’s disease would be associated with worse cognition many years before the diagnosis is made, whereas individuals with stroke may experience cognitive impairment at the time of diagnosis, as this reflects an index event of brain injury. We also compare the magnitude of cognitive deficit across these neurological conditions to provide a better understanding which may inform clinical decision-making on risk reduction strategies and long-term management.

## Methods

This study examines the UK Biobank, a population-based cohort of over 500⍰000 participants aged 38-72 years who underwent physiological measurements, cognitive testing and provided biological samples at one of 22 centres across the UK between 2006 and 2010.^20^ A subset re-attended for brain imaging between 2014 and 2020.^21^ All participants provided written informed consent. UK Biobank received approved from the North-West Multi-centre Research Ethics Committee (REC number 21/NW/0157).

The primary study objective was to investigate cognition across individuals with ischaemic stroke, focal epilepsy, Parkinson’s disease, multiple sclerosis, motor neurone disease and migraine. These diagnoses were chosen as common, acquired neurological conditions that may present to a general neurology clinic. Established cases at study baseline assessment (**post-diagnosis cases**) as well as **new diagnoses** during longitudinal follow-up were identified from hospital inpatient records, coded using the International Classification of Diseases (ICD) ICD-9 and ICD-10 codes, or from death register linkage data as an underlying or contributory cause (UK Biobank codes are found in Table S1). Secondary analysis considered all-cause dementia diagnosed after baseline assessment, which provided a benchmark with an expected cognitive decline following diagnosis, and sub-types including Alzheimer’s disease, vascular dementia and frontotemporal dementia. There were few cases of dementia at baseline assessment (N = 120), which were excluded due to the direct association with cognitive impairment. Additional exclusion criteria included other neurological conditions such as history of CNS infection, encephalitis, meningitis, haemorrhagic stroke, genetic epilepsies, previous subdural or subarachnoid haemorrhage.

### Cognitive testing

We analysed data from five computer-based cognitive tasks of working memory or speed of processing as in a previous reports focusing on executive function in the UK Biobank population,^22,23^ and used the first available timepoint data. The tests were a pairs-matching and snap reaction time, trail-making, tower-rearranging, and symbol-digit substitution tasks, described in detail elsewhere^23^ with reliability and retest-effects previously assessed.^24^

### Pre-diagnosis and post-diagnosis cognitive profiles

Cognitive profiles were created in relation to the date of diagnosis whereby post-diagnosis cognition represented all the participants with a condition who were diagnosed prior to baseline study assessment for that individual. For example, the cognitive performance of a participant who was diagnosed with multiple sclerosis 20 years prior to entering the UK Biobank study represented a 20-year post-diagnosis cognitive score for that condition. By contrast, two-year pre-diagnosis cognition reflected cognitive scores at baseline assessment for participants who would (subsequently) be diagnosed with the condition two years later during the study follow-up period.

### Main covariates

All full models were adjusted for age (continuous), sex (female vs male), education (categorised as higher [college or university degree or other professional qualification], upper secondary [second or final stage of secondary education], lower secondary [first stage of secondary education], vocational [work-related qualifications], or other), socioeconomic status (categories derived from Townsend deprivation index^25^ quintiles 1, 2 to 4, and 5).

### Brain imaging variables

Magnetic resonance imaging (MRI) data were acquired on a Skyra 3T scanner (Siemens; Munich, Germany) including high-resolution, T1-weighted, three-dimensional magnetisation-prepared gradient echo structural images and T2-weighted fluid-attenuated inversion recovery images. Full imaging protocols and processing pipeline have been previously described.^26^ We examined a global brain measure of total grey matter volume, useful to assess wide-spread change, and the total hippocampal volume which is related to cognition and relevant for several of the diseases such as epilepsy^27–29^ and stroke.^30^ Median absolute deviation was used to exclude outliers, and volumes were adjusted for potentially confounding baseline measures of age, age squared, head size, and imaging site.^26^

### Statistical analysis

Confirmatory factor analysis (CFA) was performed on cognitive variables to produce a continuous, summary latent measure of working memory and reaction time which we termed “Executive Function” for simplicity (this method has been previously described in detail).^23,31^ Estimating a latent variable has the methodological advantage of controlling measurement error that can artificially reduce the relationship between measured variables in standard univariate analyses.^32^ Missing cognitive data were estimated using full information maximum likelihood, which gives unbiased parameter estimates and standard errors.

The relationship between Executive Function across age was first examined for each group - ischaemic stroke, focal epilepsy, Parkinson’s disease, multiple sclerosis, motor neurone disease and migraine. We then controlled for age in two different ways for robustness: using a residual-based, sliding window approach with fixed age-quantile widths moved along the age distribution (using a 20% gaussian kernel and described previously,^23,33,34^ code: conditionalPlot.m available: https://osf.io/vmabg/). This has the methodological advantage of modelling non-linearities in the data and is not constrained by a pre-specified model. The second method included age as a covariate in a general linear model along with other baseline characteristics. The difference in Executive Function between pre-diagnosis cases, post-diagnosis cases and the control group was compared using ANOVA with post-hoc Tukey analysis to account for pairwise or multiple comparisons, p-values were two-sided with statistical significance set at p<0·05 for all analyses. Brain measures were analysed using the same methods.

The time course of cognitive scores and brain measures were visualised using the same sliding window, model-free approach with fixed quantile widths of time in relation to diagnosis. We compared this data with age-residualised cognitive scores from the control group. Motor neurone disease was not included in this visualisation due to low sample size but was analysed when comparing pre- and post-diagnosis Executive Function and brain imaging volumes. For the main exposures and covariates, there were less than 3% missing or not known data and complete case analysis was applied. New diagnoses were included if recorded between baseline until the date of first diagnosis, death, loss to follow-up, or the last surveyed hospital admission date (March 31, 2021, for England and Scotland and Feb 28, 2018, for Wales), whichever came first. These censoring dates were recommended by UK Biobank as the data was estimated to be over 90% complete in England, Scotland, and Wales. Analyses were done in Matlab R2018a or in R version 4.0.3 using the Lavaan^35^ or survival package.

## Results

The UK Biobank cohort comprised 502,536 participants at baseline. After excluding those who did not meet the inclusion criteria (N = 7216), our study included 497,252 individuals (flowchart in **Figure S1**). Overall, participants had a mean (SD) age of 57.5 (8.1) years and 54.5% were female. At study baseline, there were 6,352 with ischaemic stroke, 4,247 participants with epilepsy, 873 with Parkinson’s disease, 1,861 with multiple sclerosis, 70 participants diagnosed with motor neurone disease and 14,588 with migraine. The median time-since-diagnosis to baseline study assessment in these post-diagnosis cases ranged from 27.6 years in individuals with migraine to 3.3 years in individuals with MND.

Over 5,826,307 total follow-up years (median 12.0, interquartile range 11.2-12.7 years), there were 6,403 participants with a new diagnosis of ischaemic stroke, 2,511 participants with focal epilepsy, 2,442 with Parkinson’s disease, 454 with multiple sclerosis, 489 with motor neurone disease and 3,666 with migraine. The median time-to-diagnosis was 7.4 years from baseline study assessment in these ‘pre-diagnosis’ cases across all conditions. Individual condition demographic information can be found in Table 1 while histograms of the diagnosis dates in relation to baseline study assessment are shown in **Figure S2**.

**Table 1.**
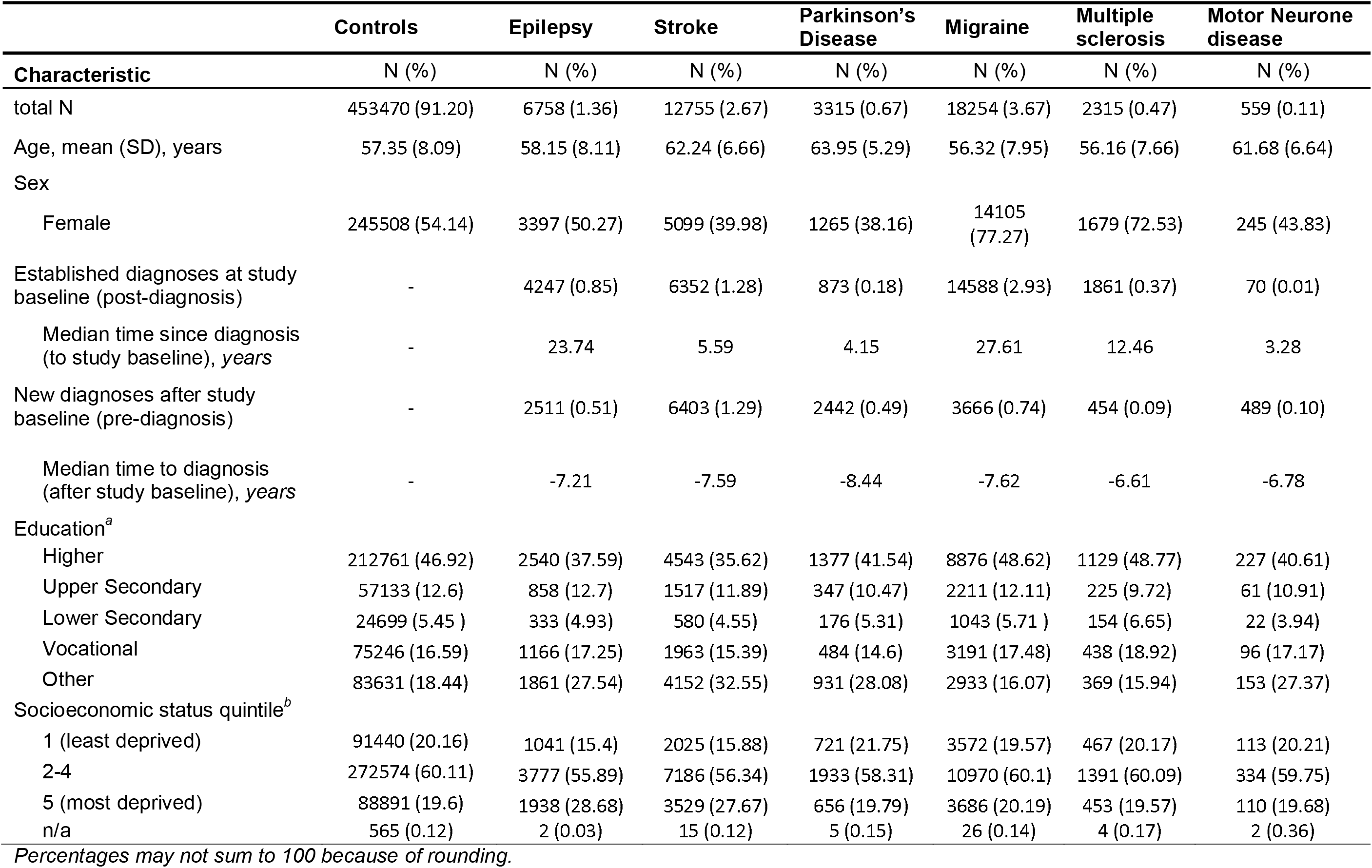
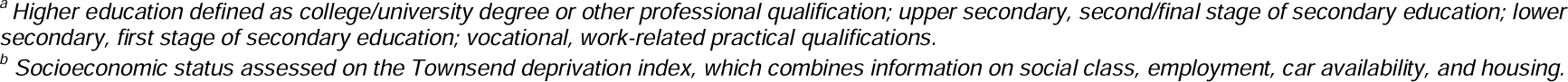
Baseline characteristics across different study sub-groups.

|  | **Controls** | **Epilepsy** | **Stroke** | **Parkinson's Disease** | **Migraine** | **Multiple sclerosis** | **Motor Neurone disease** |
| --- | --- | --- | --- | --- | --- | --- | --- |
| **Characteristic** | **N (%)** | **N (%)** | **N (%)** | **N (%)** | **N (%)** | **N (%)** | **N (%)** |
| total N | 453470 (91.20) | 6758 (1.36) | 12755 (2.67) | 3315 (0.67) | 18254 (3.67) | 2315 (0.47) | 559 (0.11) |
| Age, mean (SD), years | 57.35 (8.09) | 58.15 (8.11) | 62.24 (6.66) | 63.95 (5.29) | 56.32 (7.95) | 56.16 (7.66) | 61.68 (6.64) |
| Sex |  |  |  |  |  |  |  |
| Female | 245508 (54.14) | 3397 (50.27) | 5099 (39.98) | 1265 (38.16) | 14105 (77.27) | 1679 (72.53) | 245 (43.83) |
| Established diagnoses at study baseline (post-diagnosis) | - | 4247 (0.85) | 6352 (1.28) | 873 (0.18) | 14588 (2.93) | 1861 (0.37) | 70 (0.01) |
| Median time since diagnosis (to study baseline), years | - | 23.74 | 5.59 | 4.15 | 27.61 | 12.46 | 3.28 |
| New diagnoses after study baseline (pre-diagnosis) | - | 2511 (0.51) | 6403 (1.29) | 2442 (0.49) | 3666 (0.74) | 454 (0.09) | 489 (0.10) |
| Median time to diagnosis (after study baseline), years | - | -7.21 | -7.59 | -8.44 | -7.62 | -6.61 | -6.78 |
| Education<sup>a</sup> |  |  |  |  |  |  |  |
| Higher | 212761 (46.92) | 2540 (37.59) | 4543 (35.62) | 1377 (41.54) | 8876 (48.62) | 1129 (48.77) | 227 (40.61) |
| Upper Secondary | 57133 (12.6) | 858 (12.7) | 1517 (11.89) | 347 (10.47) | 2211 (12.11) | 225 (9.72) | 61 (10.91) |
| Lower Secondary | 24699 (5.45 ) | 333 (4.93) | 580 (4.55) | 176 (5.31) | 1043 (5.71 ) | 154 (6.65) | 22 (3.94) |
| Vocational | 75246 (16.59) | 1166 (17.25) | 1963 (15.39) | 484 (14.6) | 3191 (17.48) | 438 (18.92) | 96 (17.17) |
| Other | 83631 (18.44) | 1861 (27.54) | 4152 (32.55) | 931 (28.08) | 2933 (16.07) | 369 (15.94) | 153 (27.37) |
| Socioeconomic status quintile<sup>b</sup> |  |  |  |  |  |  |  |
| 1 (least deprived) | 91440 (20.16) | 1041 (15.4) | 2025 (15.88) | 721 (21.75) | 3572 (19.57) | 467 (20.17) | 113 (20.21) |
| 2-4 | 272574 (60.11) | 3777 (55.89) | 7186 (56.34) | 1933 (58.31) | 10970 (60.1) | 1391 (60.09) | 334 (59.75) |
| 5 (most deprived) | 88891 (19.6) | 1938 (28.68) | 3529 (27.67) | 656 (19.79) | 3686 (20.19) | 453 (19.57) | 110 (19.68) |
| n/a | 565 (0.12) | 2 (0.03) | 15 (0.12) | 5 (0.15) | 26 (0.14) | 4 (0.17) | 2 (0.36) |

*Percentages may not sum to 100 because of rounding.*
<sup>a</sup> Higher education defined as college/university degree or other professional qualification; upper secondary, second/final stage of secondary education; lower secondary, first stage of secondary education; vocational, work-related practical qualifications.
<sup>b</sup> Socioeconomic status assessed on the Townsend deprivation index, which combines information on social class, employment, car availability, and housing.

### Pre-diagnosis and post-diagnosis Executive Function across neurological conditions

A continuous cognitive function latent variable of Executive Function was estimated from five cognitive tasks of working memory or speed of processing (model and fit indices shown in **Figure S3**). Age is often the most important confounder when considering cognition and, consistent with previous findings,^23,31^ Executive Function declined uniformly with age among all groups (**Figure S4**). We therefore controlled for age using a residual-based method. Average Executive Function scores were lower in individuals with an established diagnosis (post-diagnosis) compared to individuals before they were diagnosed with a neurological condition (pre-diagnosis), apart from in those with migraine and motor neurone disease (**Figure 1, Figure S5** shows individual condition plots and **Table S2** with post-hoc Tukey analysis results, controlling for multiple comparison testing). Further, pre-diagnosis Executive Function scores were significantly lower than controls for all conditions apart from motor neurone disease. For robustness, we used a separate general linear model to corroborate these findings and control for other covariates including sex, level of education and socioeconomic status. Similar patterns of differences were observed when comparing pre-diagnosis and post-diagnosis Executive Function scores with controls (**Table S3**).

**Figure 1.**
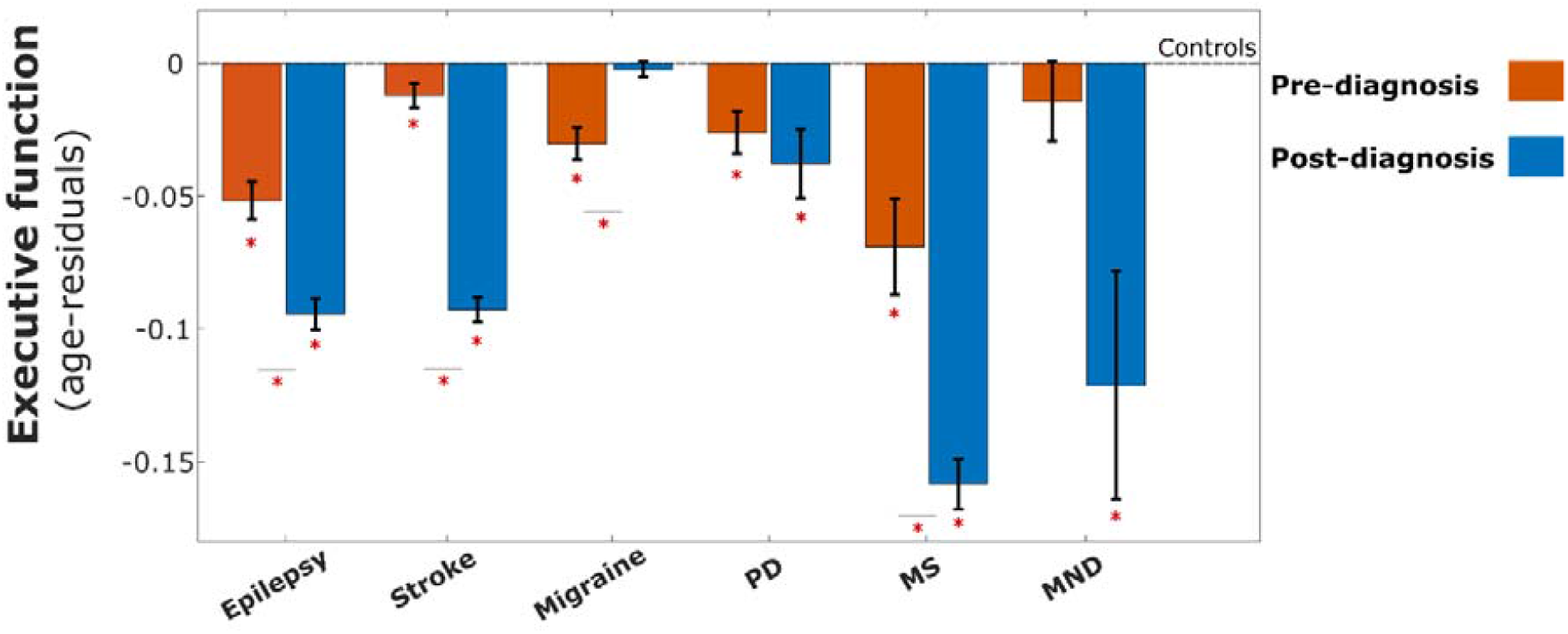
Pre-diagnosis and post-diagnosis Executive Function across different neurological conditions. Prior to being diagnosed, participants with epilepsy, stroke, migraine, Parkinson’s disease and multiple sclerosis have a lower Executive Function (z-scored values) compared to controls, with a median time-to-diagnosis of 7.4 years. Dotted baseline represents level of Executive Function in controls. Across all neurological conditions, apart from migraine and motor neurone disease, Executive function was lower in individuals with an established diagnosis (post-diagnosis cognition) compared to participants who would be diagnosed during the follow-up period of the study. Participants with migraine had a higher post-diagnosis Executive Function compared to pre-diagnosis participants. Error bars denote standard error. Mean executive function for each condition was compared with control group, asterisk (*) below each bar denotes significant difference p<0.05, while within-condition pre-diagnosis vs. post-diagnosis executive function was compared and represented by an asterisk across condition bars using a post-hoc Tukey analysis to account for multiple comparison. *PD – Parkinson’s disease, MS – multiple sclerosis, MND – motor neurone disease*

### Modelling cognitive trajectories across time in relation to diagnosis

Using a non-linear approach, Figure 2 shows the age-residual Executive Function scores leading up to and after diagnosis for each neurological condition (**Figure 2**). The upper limit of this time course reflects participants who were diagnosed many years before entering the UK Biobank study and this was different for each neurological condition. The lower limit of these cognitive trajectories represents individuals who were diagnosed during the follow-up period of the UK Biobank study (up to 15 years follow-up after baseline assessment).

**Figure 2.**
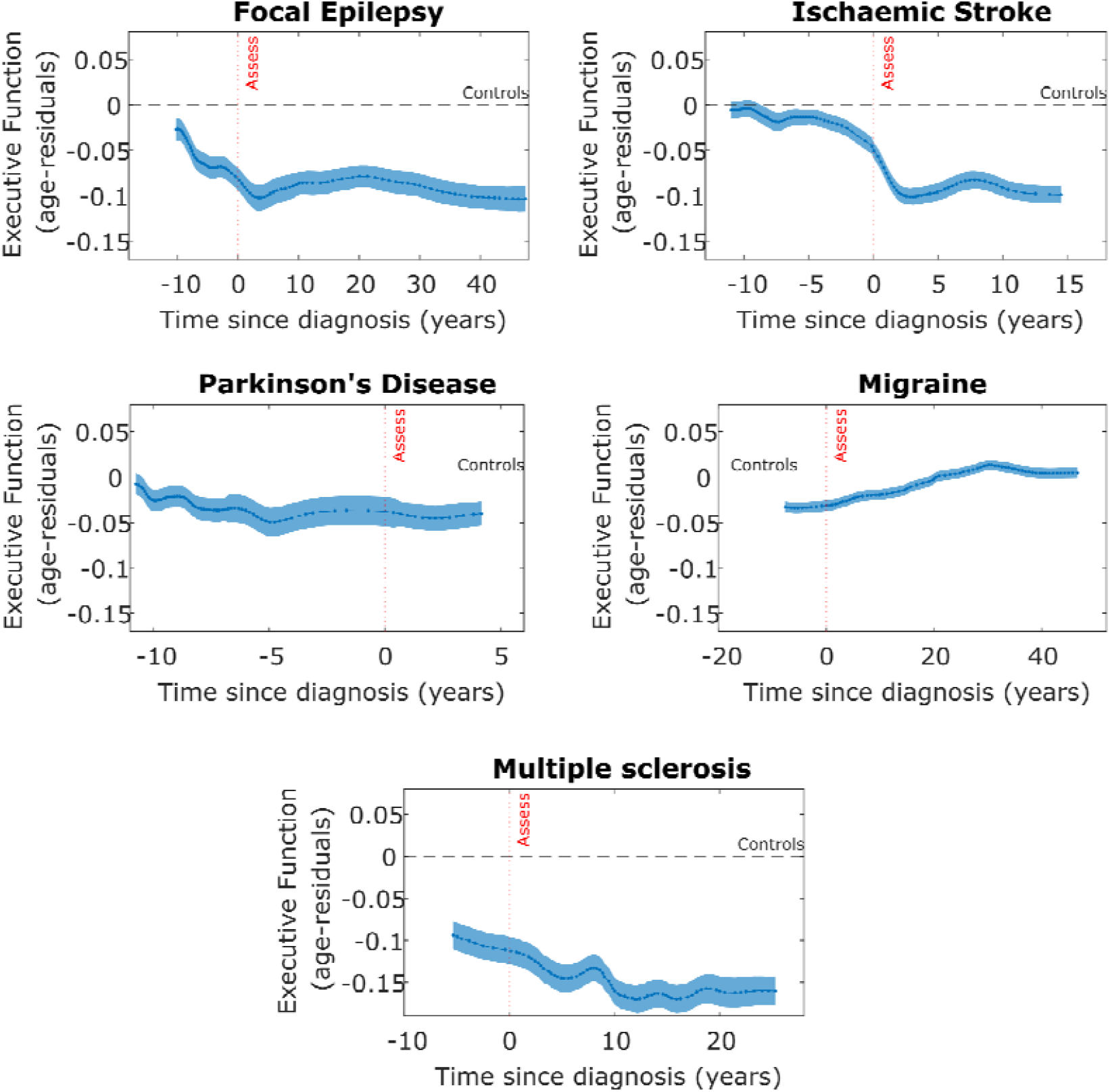
Cognitive profile across time in relation to the diagnosis of disease across different neurological conditions. Executive Function (age-residuals, z-scored values) in relation to time of diagnosis across participants with different neurological conditions represents a pre- and post-diagnosis cognitive trajectory. Black dotted line indicates the level of Executive Function in controls. Red dotted line indicates the study assessment time. Negative time values reflect individuals who *will develop* the disease (pre-diagnosis cases) during the study follow-up period. Participants with focal epilepsy, Parkinson’s disease and multiple sclerosis show declining cognitive scores prior to diagnosis at a group level. Cognitive scores plateau after diagnosis in several conditions, including focal epilepsy, and further declines in multiple sclerosis. Executive Function in participants in migraine improves following diagnosis to a level similar to controls.

This approach identified disease-specific patterns in cognitive profiles at the group level. Executive Function gradually declined in participants with focal epilepsy over 10 years before the diagnosis was made whereas participants with stroke showed Executive Function scores similar to controls up until a few years before their event, then a modest worsening followed by a substantial decline at diagnosis. Participants with multiple sclerosis had comparatively worse Executive Function compared to all other neurological conditions assessed, with a gradual decline pre-diagnosis which continued post-diagnosis. By contrast, participants with migraine had lower Executive Function before their diagnosis which improved to baseline control level after the diagnosis was made. An initial plateau in Executive Function was observed in individuals with epilepsy and stroke following the diagnosis with a small improvement between 2-10 years observed in stroke but not to the level of controls. Individuals with Parkinson’s disease showed a constant lower-than-control level of Executive Function 10 years before their diagnosis. For motor neurone disease, there was mainly data available leading up to diagnosis with few established cases at study baseline. There was worse post-diagnosis Executive Function in individuals with motor neurone disease although this was associated with large standard error due to smaller sample size.

### Pre-diagnosis cognitive profile in dementia and sub-types

There were 5963 participants who developed all-cause dementia during the follow-up period of the study. Executive Function declined progressively in individuals who were closer to receiving the diagnosis (**Figure S6**). This pattern was similar when considering those who were diagnosed with Alzheimer’s disease (2420 participants), vascular dementia (1019 participants) and other dementias (2325 participants) but was not observed in participants with frontotemporal dementia (199 participants).

### Total grey matter pre-diagnosis and post-diagnosis across neurological conditions

A subset of the UK Biobank cohort underwent MR brain imaging (N=42,468) following baseline study assessment (range 3.8 to 13.8 years after, median 9.2 years). Since imaging was obtained later, the median time-to-diagnosis from imaging acquisition was lower at 1.7 years than cognitive assessment (**Table 2** details the breakdown for each neurological condition). We used the same approach in modelling the pre-and post-diagnosis time-course for total grey matter, adjusted for age, across each neurological condition (time-course limited to 20 years post-diagnosis for visualisation in **Figure 3**). Total grey matter volume was lower than controls in individuals who have had an ischaemic stroke and those with multiple sclerosis and Parkinson’s disease, both pre- and post-diagnosis (general linear model showing statistical differences in **Table S4**). Participants with epilepsy showed decreasing total grey matter volume post-diagnosis but have volumes similar to controls leading up to the diagnosis while total grey matter volume in participants in migraine are comparable to controls before and after the diagnosis. The results for participants with motor neurone disease were not visualised because only 19 participants had imaging data.

**Table 2.** Number of participants in the imaging sub-cohort and temporal relation to diagnosis across different neurological conditions.

|  | **Controls** | **Epilepsy** | **Stroke** | **Parkinson's Disease** | **Migraine** | **Multiple sclerosis** | **Motor Neurone disease** |
| --- | --- | --- | --- | --- | --- | --- | --- |
|  | N (%) | N (%) | N (%) | N (%) | N (%) | N (%) | N (%) |
| **Characteristic** |  |  |  |  |  |  |  |
| Total N (imaging cohort) | 42468 (93.57) | 370 (0.87) | 636 (1.50) | 94 (0.22) | 1973 (4.65) | 170 (0.40) | 19 (0.04) |
| Established diagnoses at brain scan visit (post-diagnosis) | - | 321 (0.76) | 468 (1.10) | 56 (0.13) | 1852 (4.36) | 155 (0.36) | 3 (0.07) |
| Median time since diagnosis (to brain scan), years | - | 27.36 | 10.87 | 6.25 | 33.68 | 17.00 | 2.62 |
| New diagnoses after brain scan visit (pre-diagnosis) | - | 49 (0.12) | 168 (0.40) | 38 (0.09) | 121 (0.28) | 15 (0.04) | 16 (0.04) |
| Median time to diagnosis (after brain scan), years | - | -1.80 | -1.60 | -2.26 | -1.56 | -1.39 | -1.97 |

*Percentages may not sum to 100 because of rounding.*

**Figure 3.**
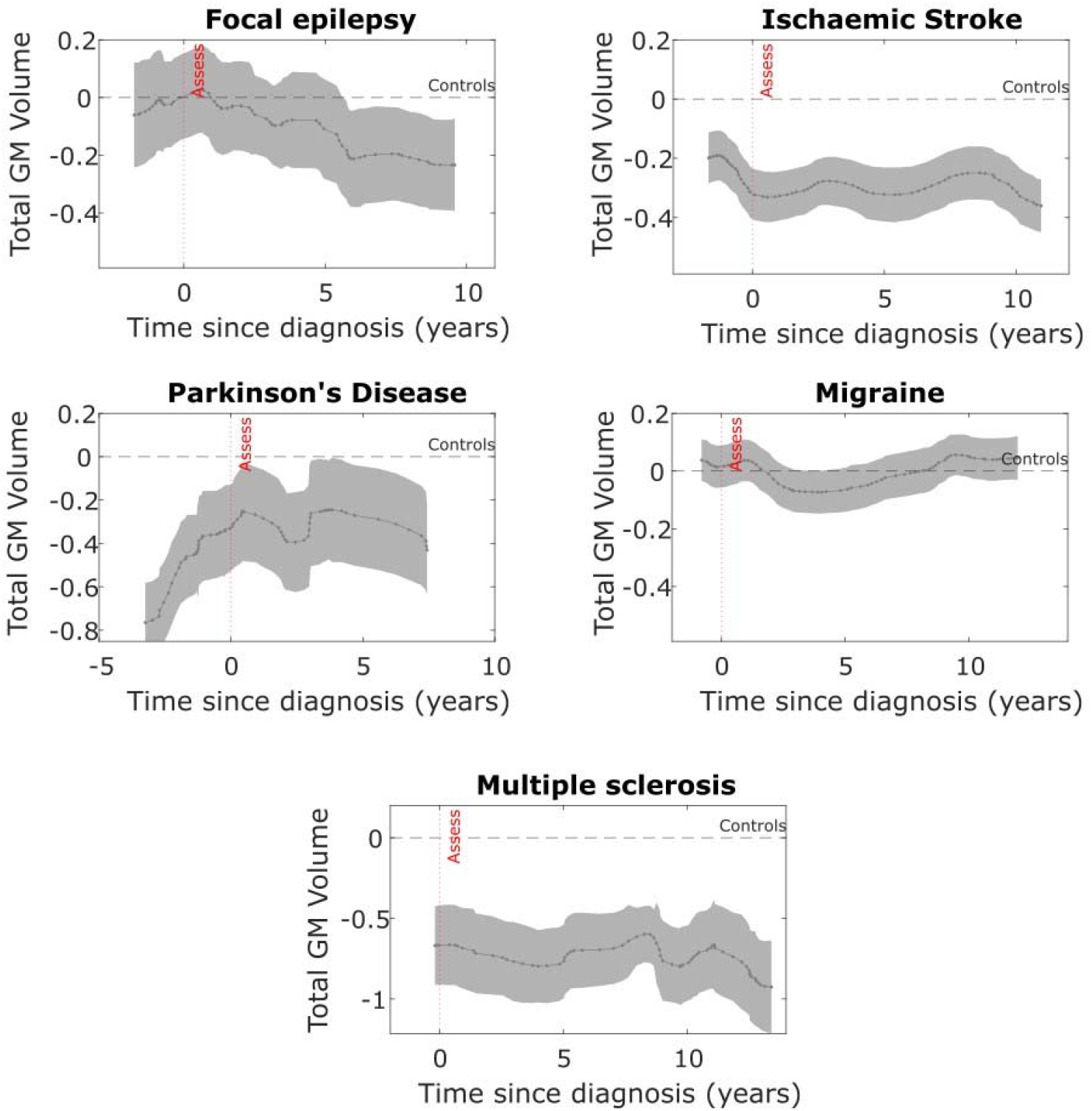
Total grey matter volume across time in relation to diagnosis across different neurological conditions. Total grey matter (GM) volume (age-residuals, z-scored) in relation to time of diagnosis across different neurological conditions. Black dotted line indicates the total grey matter volume in controls. Red dotted line indicates time of the brain scan event. Negative time values reflect individuals who will develop the disease (pre-diagnosis cases) after the brain scan has been performed. At a group level, total grey matter volume in participants with multiple sclerosis, Parkinson’s disease and a history of ischaemic stroke are lower than controls before and after the diagnosis has been made. Participants with migraine have a similar age-corrected total grey matter volume throughout the time course. In participants with epilepsy, post-diagnosis total grey matter volume decreases more steeply than expected for age in participants who have had the disease longer.

The same pre- and post-diagnosis imaging analysis was performed for total hippocampal volume which showed lower pre- and post-diagnosis volume in participants with multiple sclerosis only, while ischaemic stroke and focal epilepsy was associated with lower post-diagnosis hippocampal volume only (**Table S5**), suggesting that different disorders may be weighted towards affecting different brain areas.

## Discussion

This study examined pre- and post-diagnosis cognitive profiles in large groups of individuals with ischaemic stroke, focal epilepsy, Parkinson’s disease, multiple sclerosis and motor neurone disease. By leveraging cross-sectional cognitive data at baseline study assessment along with time-to-diagnosis (in the future) and time-since-diagnosis (from the past), we analysed group-level cognitive ‘trajectories’ with profiles up to 15 years pre-diagnosis. Such trajectories have not been clearly described previously for several of these conditions. Our residual-based, sliding-window analysis allows non-linearities to emerge from the data which more closely approximates real-world disease characteristics rather than constraining analysis to linear or quadratic functions. All conditions, apart from motor neurone disease, showed a measurable cognitive deficit several years before diagnosis compared to controls, with multiple sclerosis having the lowest cognitive score across all conditions.

Our findings offer several potential disease-specific insights. Focal epilepsy has been closely linked to cognitive impairment^36,37^, however, this has been difficult to dissociate from cognitive side-effects of anti-seizure medication.^15^ In the present study, impaired cognition was evident over a decade prior to the diagnosis. This may reflect ongoing sub-clinical epileptic activity^38^ or associated neuropathology^39^ before two overt seizures occur which are required for diagnosis.^40^ Cognitive scores plateaued within two years following focal epilepsy diagnosis which may be due to treatment stability before further decline after 20 years. Pre-diagnosis cognitive deficits observed with multiple sclerosis may reflect emerging research on early B-cell related, cortical pathology that can occur before the characteristic white matter demyelinating lesions.^16,41^

A sharp decline in cognition was observed around the diagnosis of ischaemic stroke, likely reflecting the index stroke event, while a modest reduction in cognitive scores in the few years prior may indicate underlying small vessel disease related to vascular risk factors.^31,42,43^ Interestingly, a small increase in cognitive scores between up to eight years following stroke diagnosis was observed which warrants further analysis and may relate to factors of stroke rehabilitation and brain recovery processes such as neuroplasticity.^44–46^ By contrast, individuals with migraine had lower cognitive scores leading up to the diagnosis that improved post-diagnosis back to a level similar to controls. This may represent generalised cognitive dysfunction of attention and working memory seen with migraine,^47,48^ commonly described as a ‘brain fog’, but with a potentially beneficial treatment effect following diagnosis.^49^ Importantly, the lack of ongoing cognitive decline is in line with the absence of underlying progressive or degenerative pathology in migraine.^50,51^

The neuroimaging findings suggest that reduced global brain volume can be detected in participants with ischaemic stroke, Parkinson’s disease and multiple sclerosis several years before the diagnosis. While this is consistent with the neurodegenerative and neuroinflammatory processes in Parkinson’s disease^52^ and multiple sclerosis,^41^ respectively, this has not been clearly reported before and will benefit from a closer examination of different brain regions involved in each condition.

Several further observations can be made for each individual condition, but it is important to consider our findings in the context of the study limitations. Firstly, this is a broad overview into each condition as disease-specific characteristics have not been accounted for, such the location of demyelinating lesions in participants with multiple sclerosis. Medications may be more important for some of the studied diseases, for example, we have previously shown that the number of anti-seizure medications correlates to worse cognition likely as a marker of seizure severity rather than medication side effect.^22^ In-depth analysis in each disease is the subject of ongoing investigation. However, in our view, this general approach provides a useful and novel comparison across these conditions.

Crucially, due to the observational nature of this cohort, any associations or findings cannot be taken as causal and potential mechanistic explanations are hypothetical. Diagnosis information was based on hospital records or death certificate data and therefore delayed or incorrect diagnoses may affect the results. Nonetheless, early changes were observed up to 20 years before diagnosis for several of these conditions. Electronic records may be more accurate for certain conditions, such as ischaemic stroke, when there is usually a clear event that brings an individual to hospital compared to neurodegenerative conditions with a longer diagnostic journey. However, the sample size of each sub-group is an order of magnitude larger than most separate studies of each condition and will help identify small effects within the data. The cross-sectional nature of the data means that these cognitive and brain models likely capture a combination of disease trajectory over time and cohort-specific characteristics. This includes a selection bias that reflects the relatively healthy nature of the UK Biobank^53^ and therefore the effects described may underestimate the magnitude of cognitive changes should a more representative sample of each condition were considered.

## Conclusion

Analysis of cognitive and brain trajectories can offer novel disease-specific insights and improve understanding of underlying pathological mechanisms. Cognitive deficits were identified over a decade before diagnosis in conditions such as epilepsy and multiple sclerosis while cognition post-diagnosis improved to a healthy control baseline in individuals with migraine but declined across other conditions. This non-linear approach using cross-sectional cognitive data with time-to-diagnosis and time-since-diagnosis is made possible with the large cohort of the UK Biobank and is a cost-effective and useful method to understand disease time-courses compared to longitudinal studies. These findings may help prioritise risk management of individual diseases and may inform screening, clinical diagnosis and long-term decision-making in the future.

## Data Availability

The data analysed during the current study are available from the UK Biobank https://www.ukbiobank.ac.uk/researchers/. The variables used are detailed in Supplementary Table 1. This research was conducted using the UK Biobank resource under application 9462.

## Acknowledgments

This work was funded by the Wellcome Trust (Wellcome Trust PhD clinical fellowship to XYT and Wellcome Trust Principal Research Fellowship to MH [206330/Z/17/Z]) and MRC (Clinician Scientist Fellowship to S.M. [MR/P00878/X]) and by the Oxford NIHR Biomedical Research Centre.

## Author contributions

Design, analysis, manuscript writing, critical revisions (XYT); design, statistical analysis and critical revisions of the manuscript (ST, DL, CvD, MH, SM)

## References

1. Sperling RA, Aisen PS, Beckett LA, et al. Toward defining the preclinical stages of Alzheimer’s disease: Recommendations from the National Institute on Aging-Alzheimer’s Association workgroups on diagnostic guidelines for Alzheimer’s disease. Alzheimers Dement. 2011;7(3):280. doi:10.1016/J.JALZ.2011.03.003

2. Hardy J, Selkoe DJ. The amyloid hypothesis of Alzheimer’s disease: Progress and problems on the road to therapeutics. Science (1979). 2002;297(5580):353–356. doi:10.1126/SCIENCE.1072994/ASSET/C4ED1918-4F2B-4649-BA69-684D0145F32B/ASSETS/GRAPHIC/SE2820694002.JPEG

3. Guo T, Zhang D, Zeng Y, Huang TY, Xu H, Zhao Y. Molecular and cellular mechanisms underlying the pathogenesis of Alzheimer’s disease. Molecular Neurodegeneration 2020 15:1. 2020;15(1):1–37. doi:10.1186/S13024-020-00391-7

4. Howard R, McShane R, Lindesay J, et al. Donepezil and Memantine for Moderate-to-Severe Alzheimer’s Disease. New England Journal of Medicine. 2012;366(10):893–903. doi:10.1056/NEJMOA1106668/SUPPL_FILE/NEJMOA1106668_DISCLOSURES.PDF

5. van Dyck CH, Swanson CJ, Aisen P, et al. Lecanemab in Early Alzheimer’s Disease. N Engl J Med. Published online November 29, 2022. doi:10.1056/NEJMOA2212948/SUPPL_FILE/NEJMOA2212948_APPENDIX.PDF

6. Mintun MA, Lo AC, Evans CD, et al. Donanemab in Early Alzheimer’s Disease. 101056/NEJMoa2100708. 2021;384(18):1691–1704. doi:10.1056/NEJMOA2100708

7. Mattsson-Carlgren N, Salvadó G, Ashton NJ, et al. Prediction of Longitudinal Cognitive Decline in Preclinical Alzheimer Disease Using Plasma Biomarkers. JAMA Neurol. 2023;80(4):360–369. doi:10.1001/JAMANEUROL.2022.5272

8. Almkvist O, Rodriguez-Vieitez E, Thordardottir S, et al. Longitudinal cognitive decline in autosomal-dominant Alzheimer’s disease varies with mutations in APP and PSEN1 genes. Neurobiol Aging. 2019;82:40–47. doi:10.1016/J.NEUROBIOLAGING.2019.06.010

9. Bateman RJ, Xiong C, Benzinger TLS, et al. Clinical and Biomarker Changes in Dominantly Inherited Alzheimer’s Disease. New England Journal of Medicine. 2012;367(9):795–804. doi:10.1056/NEJMOA1202753/SUPPL_FILE/NEJMOA1202753_DISCLOSURES.PDF

10. Petersen RC, Smith GE, Waring SC, Ivnik RJ, Tangalos EG, Kokmen E. Mild Cognitive Impairment: Clinical Characterization and Outcome. Arch Neurol. 1999;56(3):303–308. doi:10.1001/ARCHNEUR.56.3.303

11. Insel PS, Weiner M, Scott MacKin R, et al. Determining clinically meaningful decline in preclinical Alzheimer disease. Neurology. 2019;93(4):e322. doi:10.1212/WNL.0000000000007831

12. Sun JH, Tan L, Yu JT. Post-stroke cognitive impairment: epidemiology, mechanisms and management. Ann Transl Med. 2014;2(8):80. doi:10.3978/J.ISSN.2305-5839.2014.08.05

13. Rasquin SMC, Verhey FRJ, Van Oostenbrugge RJ, Lousberg R, Lodder J. Demographic and CT scan features related to cognitive impairment in the first year after stroke. J Neurol Neurosurg Psychiatry. 2004;75(11):1562. doi:10.1136/JNNP.2003.024190

14. Chausson N, Olindo S, Cabre P, Saint-Vil M, Smadja D. Five-year outcome of a stroke cohort in Martinique, French West Indies: Etude Réalisée en Martinique et Centrée sur l’Incidence des Accidents vasculaires cérebraux, Part 2. Stroke. 2010;41(4):594–599. doi:10.1161/STROKEAHA.109.573402

15. Witt JA, Werhahn KJ, Krämer G, Ruckes C, Trinka E, Helmstaedter C. Cognitive-behavioral screening in elderly patients with new-onset epilepsy before treatment. Acta Neurol Scand. 2014;130(3):172–177. doi:10.1111/ANE.12260

16. Lucchinetti CF, Popescu BFG, Bunyan RF, et al. Inflammatory Cortical Demyelination in Early Multiple Sclerosis. N Engl J Med. 2011;365(23):2188. doi:10.1056/NEJMOA1100648

17. Galovic M, Van Dooren VQH, Postma TS, et al. Progressive Cortical Thinning in Patients With Focal Epilepsy. JAMA Neurol. 2019;76(10):1230–1239. doi:10.1001/JAMANEUROL.2019.1708

18. Werden E, Cumming T, Li Q, et al. Structural MRI markers of brain aging early after ischemic stroke. Neurology. 2017;89(2):116–124. doi:10.1212/WNL.0000000000004086

19. Lorefice L, Frau J, Coghe G, et al. Assessing the burden of vascular risk factors on brain atrophy in multiple sclerosis: A case-control MRI study. Mult Scler Relat Disord. 2019;27:74–78. doi:10.1016/J.MSARD.2018.10.011

20. Sudlow C, Gallacher J, Allen N, et al. UK biobank: an open access resource for identifying the causes of a wide range of complex diseases of middle and old age. PLoS Med. 2015;12(3):e1001779. doi:10.1371/journal.pmed.1001779

21. Littlejohns TJ, Holliday J, Gibson LM, et al. The UK Biobank imaging enhancement of 100,000 participants:lllrationale, data collection, management and future directions. Nature Communications 2020 11:1. 2020;11(1):1–12. doi:10.1038/s41467-020-15948-9

22. Tai XY, Torzillo E, Lyall DM, et al. Association of Dementia Risk With Focal Epilepsy and Modifiable Cardiovascular Risk Factors. JAMA Neurol. Published online March 27, 2023. doi:10.1001/JAMANEUROL.2023.0339

23. Tai XY, Chen C, Manohar S, Husain M. Impact of sleep duration on executive function and brain structure. Communications Biology 2022 5:1. 2022;5(1):1–10. doi:10.1038/s42003-022-03123-3

24. Fawns-Ritchie C, Deary I. Reliability and validity of the UK Biobank cognitive tests. Published online 2019. doi:10.1101/19002204

25. Townsend P. Deprivation*. J Soc Policy. 1987;16(2):125–146. doi:10.1017/S0047279400020341

26. Alfaro-Almagro F, Jenkinson M, Bangerter NK, et al. Image processing and Quality Control for the first 10,000 brain imaging datasets from UK Biobank. Neuroimage. 2018;166:400–424. doi:10.1016/j.neuroimage.2017.10.034

27. Dabbs K, Becker T, Jones J, Rutecki P, Seidenberg M, Hermann B. Brain structure and aging in chronic temporal lobe epilepsy. Epilepsia. 2012;53(6):1033–1043. doi:10.1111/J.1528-1167.2012.03447.X

28. Caciagli L, Bernasconi A, Wiebe S, Koepp MJ, Bernasconi N, Bernhardt BC. A meta-analysis on progressive atrophy in intractable temporal lobe epilepsy: Time is brain? Neurology. 2017;89(5):506–516. doi:10.1212/WNL.0000000000004176

29. Whelan CD, Altmann A, Botía JA, et al. Structural brain abnormalities in the common epilepsies assessed in a worldwide ENIGMA study. Brain. 2018;141(2):391. doi:10.1093/BRAIN/AWX341

30. Firbank MJ, Burton EJ, Barber R, et al. Medial temporal atrophy rather than white matter hyperintensities predict cognitive decline in stroke survivors. Neurobiol Aging. 2007;28(11):1664–1669. doi:10.1016/J.NEUROBIOLAGING.2006.07.009

31. Veldsman M, Tai XY, Nichols T, et al. Cerebrovascular risk factors impact frontoparietal network integrity and executive function in healthy ageing. Nat Commun. 2020;11(1). doi:10.1038/s41467-020-18201-5

32. McDonald RP, Bollen KA. Structural Equations with Latent Variables. J Am Stat Assoc. Published online 1990. doi:10.2307/2289630

33. Nobis L, Manohar SG, Smith SM, et al. Hippocampal volume across age: Nomograms derived from over 19,700 people in UK Biobank. Neuroimage Clin. 2019;23:101904. doi:10.1016/j.nicl.2019.101904

34. Manohar SG. Matlib: MATLAB tools for plotting, data analysis, eye tracking and experiment design (Public). Published online 2019. doi:10.17605/OSF.IO/VMABG

35. Rosseel Y. Lavaan: An R package for structural equation modeling. J Stat Softw. 2012;48. doi:10.18637/jss.v048.i02

36. Sen A, Capelli V, Husain M. Cognition and dementia in older patients with epilepsy. Brain. 2018;141(6):1592–1608. doi:10.1093/BRAIN/AWY022

37. Elger CE, Helmstaedter C, Kurthen M. Chronic epilepsy and cognition. Lancet Neurol. 2004;3(11):663–672. doi:10.1016/S1474-4422(04)00906-8

38. Holmes GL, Lenck-Santini PP. Role of interictal epileptiform abnormalities in cognitive impairment. Epilepsy Behav. 2006;8(3):504–515. doi:10.1016/j.yebeh.2005.11.014

39. Tai XY, Koepp M, Duncan JS, et al. Hyperphosphorylated tau in patients with refractory epilepsy correlates with cognitive decline: A study of temporal lobe resections. Brain. 2016;139(9). doi:10.1093/brain/aww187

40. Fisher RS, Acevedo C, Arzimanoglou A, et al. A practical clinical definition of epilepsy. Epilepsia. 2014;55(4):475–482. doi:10.1111/epi.12550

41. Bhargava P, Hartung HP, Calabresi PA. Contribution of B cells to cortical damage in multiple sclerosis. Brain. 2022;145(10):3363–3373. doi:10.1093/BRAIN/AWAC233

42. Prins ND, van Dijk EJ, den Heijer T, et al. Cerebral small-vessel disease and decline in information processing speed, executive function and memory. Brain. 2005;128(9):2034–2041. doi:10.1093/brain/awh553

43. Staekenborg SS, Su T, van Straaten ECW, et al. Behavioural and psychological symptoms in vascular dementia; differences between small- and large-vessel disease. J Neurol Neurosurg Psychiatry. 2010;81(5):547–551. doi:10.1136/jnnp.2009.187500

44. Alia C, Spalletti C, Lai S, et al. Neuroplastic Changes Following Brain Ischemia and their Contribution to Stroke Recovery: Novel Approaches in Neurorehabilitation. Front Cell Neurosci. 2017;11. doi:10.3389/FNCEL.2017.00076

45. Zhao Q, Wang X, Wang T, et al. Cognitive rehabilitation interventions after stroke: protocol for a systematic review and meta-analysis of randomized controlled trials. Syst Rev. 2021;10(1):1–9. doi:10.1186/S13643-021-01607-7/FIGURES/2

46. Niimi M, Kakuda W, Abo M. Brain Plasticity and Stroke Rehabilitation. Stroke. 2000;127(4):151–167. doi:10.1161/01.STR.31.1.223

47. de Araújo CM, Barbosa IG, Aguiar Lemos SM, Domingues RB, Teixeira AL. Cognitive impairment in migraine: A systematic review. Dement Neuropsychol. 2012;6(2):74. doi:10.1590/S1980-57642012DN06020002

48. Gil-Gouveia R, Martins IP. Cognition and Cognitive Impairment in Migraine. Curr Pain Headache Rep. 2019;23(11):1–10. doi:10.1007/S11916-019-0824-7/METRICS

49. Rist PM, Kang JH, Buring JE, Glymour MM, Grodstein F, Kurth T. Migraine and cognitive decline among women: prospective cohort study. BMJ. 2012;345(7873). doi:10.1136/BMJ.E5027

50. Islamoska S, Hansen ÅM, Wang HX, et al. Mid-To late-life migraine diagnoses and risk of dementia: A national register-based follow-up study. Journal of Headache and Pain. 2020;21(1):1–12. doi:10.1186/S10194-020-01166-7/TABLES/4

51. Hurh K, Jeong SH, Kim SH, Jang SY, Park EC, Jang SI. Increased risk of all-cause, Alzheimer’s, and vascular dementia in adults with migraine in Korea: a population-based cohort study. Journal of Headache and Pain. 2022;23(1):1–9. doi:10.1186/S10194-022-01484-Y/TABLES/3

52. Dickson DW. Parkinson’s Disease and Parkinsonism: Neuropathology. Cold Spring Harb Perspect Med. 2012;2(8). doi:10.1101/CSHPERSPECT.A009258

53. Fry A, Littlejohns TJ, Sudlow C, et al. Comparison of Sociodemographic and Health-Related Characteristics of UK Biobank Participants With Those of the General Population. Am J Epidemiol. 2017;186(9):1026–1034. doi:10.1093/aje/kwx246

