## Supplementary material for "Detection of cognitive deficits years prior to clinical diagnosis across neurological conditions"

**Table S1.** Codes used in the UK Biobank study to identify dementia and cardiometabolic condition cases and exclusion diagnoses

**Table S2.** Post-hoc comparisons examining the differences in Executive Function between controls, pre-diagnosis and postdiagnosis scores across different neurological conditions

**Table S3.** Comparing pre-diagnosis and post-diagnosis Executive Function in each neurological condition using a general linear model controlling for other baseline characteristics

**Table S4.** Comparing pre-diagnosis and post-diagnosis total grey matter volume in each neurological condition using a general linear model controlling for other baseline characteristics

**Table S5.** Comparing pre-diagnosis and post-diagnosis total hippocampal volume in each neurological condition using a general linear model controlling for other baseline characteristics

**Figure S1.** Study flowchart

**Figure S2.** Histogram of when neurological conditions were diagnosed in relation to study assessment.

**Figure S3.** Confirmatory factor analysis of computer-based cognitive tasks

**Figure S4.** Executive Function across age for different neurological conditions

**Figure S5.** Comparing pre-diagnosis and post-diagnosis Executive Function for each individual neurological condition

**Figure S6.** Pre-diagnosis cognitive profile for participants with dementia

**Table S1. Codes used in the UK Biobank study to identify dementia and cardiometabolic condition cases and exclusion diagnoses**

|  | **Algorithmically-derived** | **Self-report (includes non-cancer and treatment self report)** | **Illness code: ICD-10** | **Illness code: ICD-9** |
| --- | --- | --- | --- | --- |
| **Dementia** | **-** | 1263 | AD: F00, F00.0, F00.1, F00.2, F00.9, G30, G30.0, G30.1, G30.8, G30.9 VaD: F01, F01.0, F01.1, F01.2, F01.3, F01.8, F01.9, I67.3 FTD: F02.0, G31.0, Other codes for all-cause dementia: A81.0, F02, F02.1, F02.2, F02.3, F02.4, F02.8, F03, F05.1, F10.6, G31.1, G31.8 | AD: 331.0 VaD: 290.4 FTD: 331.1 Other codes for all-cause dementia: 290.2, 290.3, 291.2, 294.1, 331.2, 331.5 |
| **Stroke** | All-cause stroke: 42006 | 1081 | I630, I631, I632, I633, I634, I635, I636, I638, I639, | 43491 |
| **Epilepsy** | - | 1264 | G40, G400, G401, G402,  G405, G406, G407, G408, G409, G41, G410, G411, G412, G418,G419 | 34540, 34540, 34541, 34550, 34551 |
| **Migraine** |  | 1265 | G43, G430, G431, G432, G433, G438, G439 | 34690 |
| **Motor Neurone Disease (Amyotrophic lateral sclerosis)** | **-** | 1259 | G122 | 3352 |
| **Multiple Sclerosis** | **-** | 1261 | G35, G350 | **-** |
| **Infection of the central nervous system** | **-** | 1244 | **-** | **-** |
| **Encephalitis** | **-** | 1246 | **-** | **-** |
| **Meningitis** | **-** | 1247 | **-** | **-** |
|  | **-** | 1262 |  |  |
| **Head Injury** | **-** | 1266 | **-** | **-** |
| **Subdural Haematoma** | **-** | 1083 | **-** | **-** |
| **Subarachnoid Haemorrhage** | **-** | 1086 | **-** | **-** |

**Table S2. Post-hoc comparisons examining the differences in Executive Function between controls, pre-diagnosis and postdiagnosis scores across different neurological conditions**

|  | **Epilepsy** | | **Stroke** | | **Parkinson's disease** | | **Migraine** | | **Multiple Sclerosis** | | **Motor Neurone disease** | |
| --- | --- | --- | --- | --- | --- | --- | --- | --- | --- | --- | --- | --- |
|  | **F** | **p** | **F** | **p** | **F** | **p** | **F** | **p** | **F** | **p** | **F** | **p** |
| **Overall model** | 189.70 | *< .001 | 235.08 | *< .001 | 14.31 | *< .001 | 18.48 | *< .001 | 202.40 | *< .001 | 5.05 | *< .001 |
| **Post-hoc analysis** | **Mean diff (95% CI)** | **P_Tukey_** | **Mean diff (95% CI)** | **P_Tukey_** | **Mean diff (95% CI)** | **P_Tukey_** | **Mean diff (95% CI)** | **P_Tukey_** | **Mean diff (95% CI)** | **P_Tukey_** | **Mean diff (95% CI)** | **P_Tukey_** |
| **Individual groups** |  |  |  |  |  |  |  |  |  |  |  |  |
| Controls – Pre-diagnosis | 0.06 (0.04 - 0.07) | *< .001 | 0.02 (0.01 - 0.03) | *< .001 | 0.03 (0.01 - 0.05) | *< .001 | 0.03 (0.02 - 0.05) | *< .001 | 0.07 (0.03 - 0.11) | *< .001 | 0.02 (-0.02 - 0.06) | 0.5 |
| Controls – Post-diagnosis | 0.1 (0.09 - 0.11) | *< .001 | 0.1 (0.09 - 0.11) | *< .001 | 0.04 (0.01 - 0.07) | *< .001 | 0.01 (0.00 - 0.01) | 0.11 | 0.16 (0.14 - 0.18) | *< .001 | 0.12 (0.03 - 0.22) | *0.01 |
| Pre-diagnosis – Post-diagnosis | 0.04 (0.02 - 0.06) | *< .001 | 0.08 (0.07 - 0.10) | *< .001 | 0.01 (-0.02 - 0.04) | 0.68 | -0.03  (-0.04 – -0.01) | *< .001 | 0.09 (0.05 - 0.13) | *< .001 | 0.11 (0 - 0.21) | *0.05 |

*Post-hoc analysis of relationship between individual groups of controls, pre-diagnosis cognition and post-diagnosis cognition for each neurological conditions with a Tukey correction applied to account for multiple comparisons. CI- confidence interval.*

**Table S3. Comparing pre-diagnosis and post-diagnosis Executive Function in each neurological condition using a general linear model controlling for other baseline characteristics**

|  | **Epilepsy** | | | **Stroke** | | | **Parkinson's disease** | | | **Migraine** | | | **Multiple Sclerosis** | | | **Motor Neurone disease** | | |
| --- | --- | --- | --- | --- | --- | --- | --- | --- | --- | --- | --- | --- | --- | --- | --- | --- | --- | --- |
|  | **Full Model^a^** | | | **Full Model^a^** | | | **Full Model^a^** | | | **Full Model^a^** | | | **Full Model^a^** | | | **Full Model^a^** | | |
|  | **β** | **t-stat** | **p** | **β** | **t-stat** | **p** | **β** | **t-stat** | **p** | **β** | **t-stat** | **p** | **β** | **t-stat** | **p** | **β** | **t-stat** | **p** |
| **Executive function** |  |  |  |  |  |  |  |  |  |  |  |  |  |  |  |  |  |  |
| Controls |  | Baseline |  |  | Baseline |  |  | Baseline |  |  | Baseline |  |  | Baseline |  |  | Baseline |  |
| Pre-diagnosis | -0.05 | -5.89 | *< .001 | -0.01 | -2.52 | 0.01 | -0.03 | -3.46 | *< .001 | -0.02 | -2.99 | *< .001 | -0.07 | -3.98 | *< .001 | 0.00 | -0.20 | 0.84 |
| Post-diagnosis | -0.08 | -12.87 | *< .001 | -0.09 | -17.00 | *< .001 | -0.06 | -4.43 | *< .001 | 0.00 | 0.64 | 0.52 | -0.15 | -17.33 | *< .001 | -0.10 | -1.98 | 0.05 |
| **Age** | -0.01 | -171.33 | *< .001 | -0.01 | -171.51 | *< .001 | -0.01 | -170.61 | *< .001 | -0.01 | -173.64 | *< .001 | -0.01 | -170.73 | *< .001 | -0.01 | -170.67 | *< .001 |

*^a^Full model: General linear model adjusted for baseline characteristics including age, sex, education, socioeconomic status and assessment centre.* β *= unstandardised betas*

**Table S4.** **Comparing pre-diagnosis and post-diagnosis total grey matter volume in each neurological condition using a general linear model controlling for other baseline characteristics**

|  | **Epilepsy** | | | **Stroke** | | | **Parkinson's disease** | | | **Migraine** | | | **Multiple Sclerosis** | | | **Motor Neurone disease** | | |
| --- | --- | --- | --- | --- | --- | --- | --- | --- | --- | --- | --- | --- | --- | --- | --- | --- | --- | --- |
|  | **Full Model^a^** | | | **Full Model^a^** | | | **Full Model^a^** | | | **Full Model^a^** | | | **Full Model^a^** | | | **Full Model^a^** | | |
|  | **β** | **t-stat** | **p** | **β** | **t-stat** | **p** | **β** | **t-stat** | **p** | **β** | **t-stat** | **p** | **β** | **t-stat** | **p** | **β** | **t-stat** | **p** |
| **Total Grey Matter volume** |  |  |  |  |  |  |  |  |  |  |  |  |  |  |  |  |  |  |
| Controls | Baseline | | | Baseline | | | Baseline | | | Baseline | | | Baseline | | | Baseline | | |
| Pre-diagnosis | -0.05 | -0.33 | 0.738 | -0.23 | -2.64 | *0.008 | -0.60 | -3.23 | *0.001 | 0.10 | 0.97 | 0.331 | -0.64 | -2.32 | *0.020 | -0.86 | -3.10 | *0.002 |
| Post-diagnosis | -0.29 | -4.71 | *< .001 | -0.34 | -6.18 | *< .001 | -0.31 | -2.13 | 0.033 | 0.05 | 1.89 | 0.059 | -0.62 | -6.84 | *< .001 | -0.26 | -0.45 | 0.655 |

*^a^Full model: General linear model adjusted for baseline characteristics including age, sex, education, socioeconomic status and assessment centre.* β *= unstandardised betas*

**Table S5.** **Comparing pre-diagnosis and post-diagnosis total hippocampal volume in each neurological condition using a general linear model controlling for other baseline characteristics**

|  | **Epilepsy** | | | **Stroke** | | | **Parkinson's disease** | | | **Migraine** | | | **Multiple Sclerosis** | | | **Motor Neurone disease** | | |
| --- | --- | --- | --- | --- | --- | --- | --- | --- | --- | --- | --- | --- | --- | --- | --- | --- | --- | --- |
|  | **Full Model^a^** | | | **Full Model^a^** | | | **Full Model^a^** | | | **Full Model^a^** | | | **Full Model^a^** | | | **Full Model^a^** | | |
|  | **β** | **t-stat** | **p** | **β** | **t-stat** | **p** | **β** | **t-stat** | **p** | **β** | **t-stat** | **p** | **β** | **t-stat** | **p** | **β** | **t-stat** | **p** |
| **Total Hippocampal volume** |  |  |  |  |  |  |  |  |  |  |  |  |  |  |  |  |  |  |
| Controls | Baseline | | | Baseline | | | Baseline | | | Baseline | | | Baseline | | | Baseline | | |
| Pre-diagnosis | -0.178 | -1.11 | 0.26 | -0.06 | -0.69 | 0.49 | -0.37 | -2.00 | 0.05 | -0.08 | -0.80 | 0.43 | -1.1 | -3.99 | *< .001 | -0.74 | -2.68 | *0.01 |
| Post-diagnosis | -0.264 | -4.31 | *< .001 | -0.33 | -6.03 | *< .001 | 0.14 | 0.93 | 0.35 | 0.02 | 0.66 | 0.51 | -1.17 | -12.93 | *< .001 | 0.34 | 0.60 | 0.55 |

*^a^Full model: General linear model adjusted for baseline characteristics including age, sex, education, socioeconomic status and assessment centre.* β *= unstandardised betas*

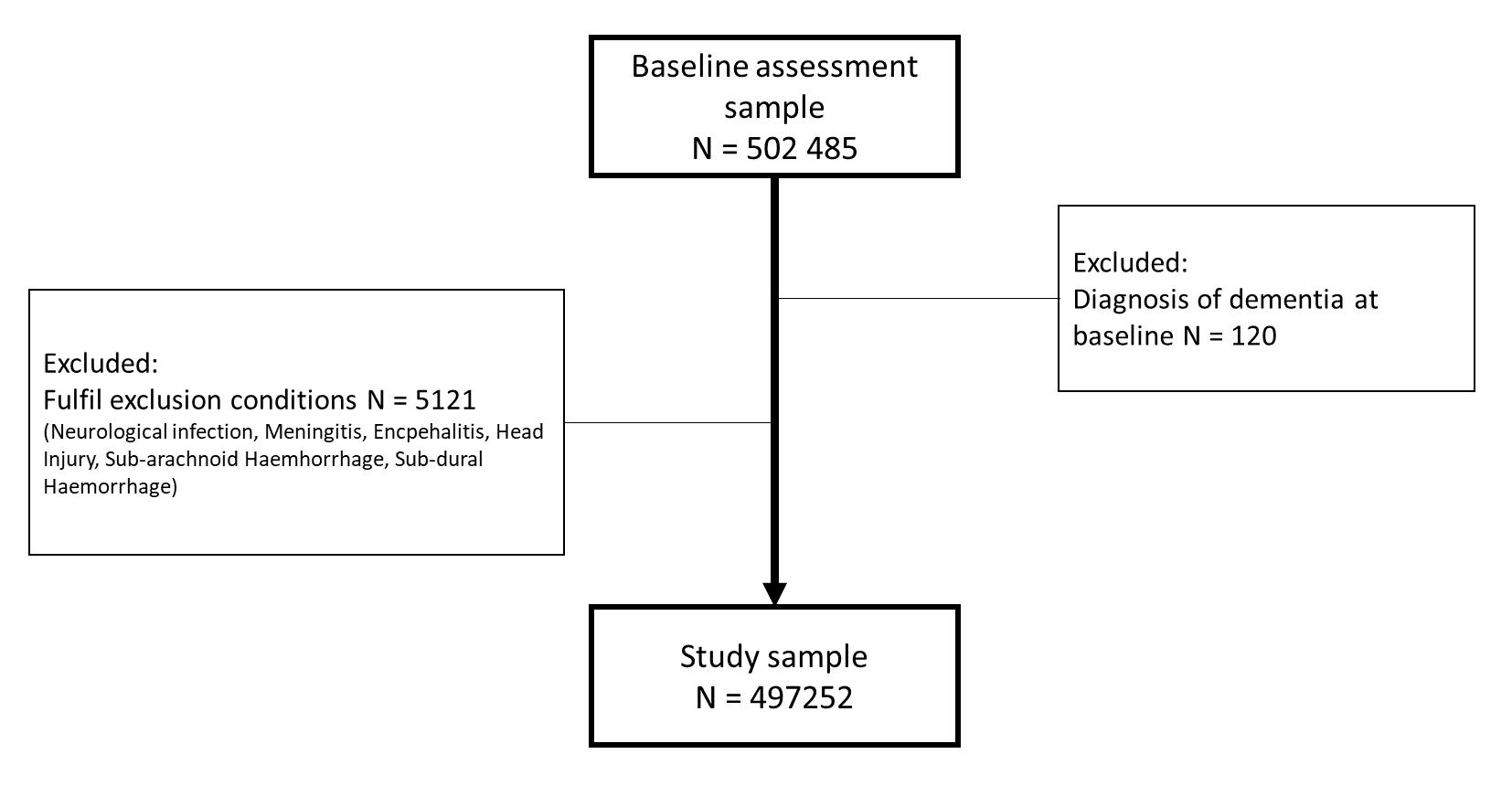

**Figure S1. Study flowchart**

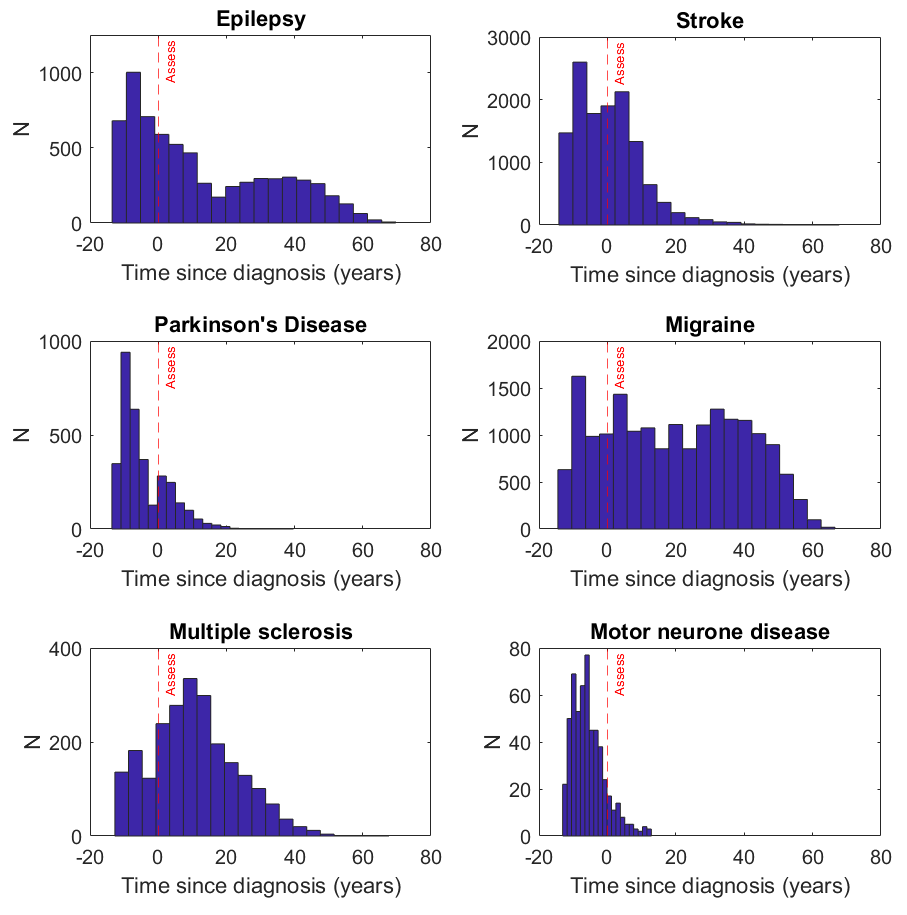

**Figure S2. Histogram of when neurological conditions were diagnosed in relation to study assessment.**

Time-course of diagnoses across different neurological conditions. Red dotted line indicates the baseline study assessment. Negative time course values indicate that the participant will be diagnosed after initial assessment, during the follow-up period of the study.

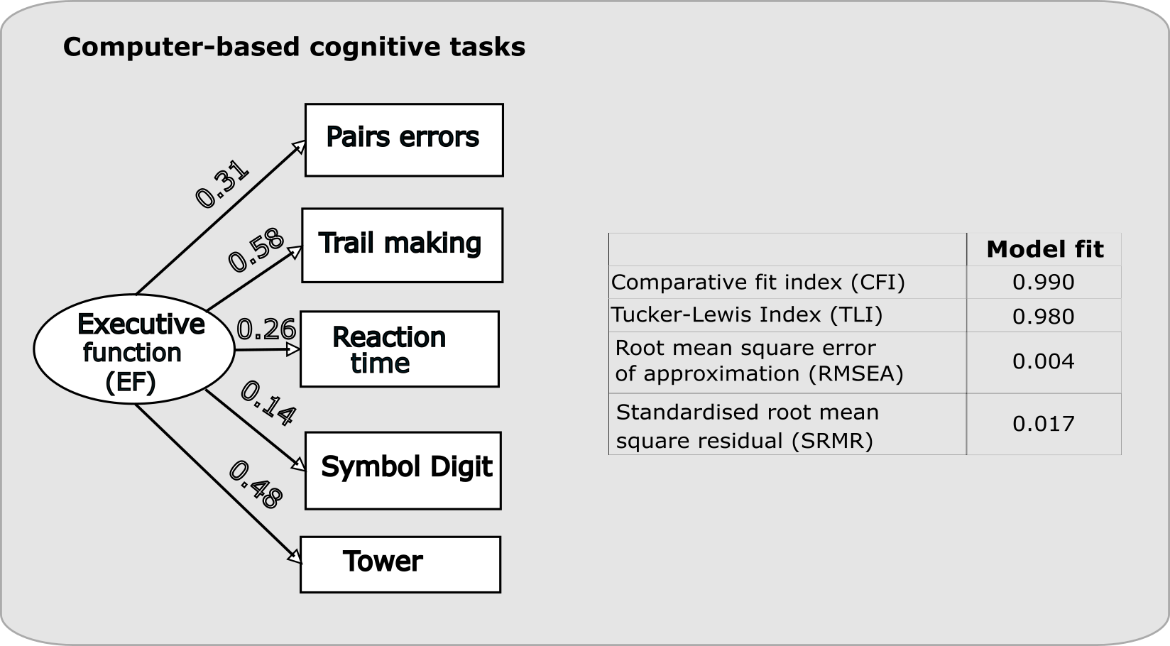

**Figure S3. Confirmatory factor analysis of computer-based cognitive tasks**

Path diagram (left) used to create a single latent variable of Executive Function (EF) using confirmatory factor analysis. Fit indices shown on the right.

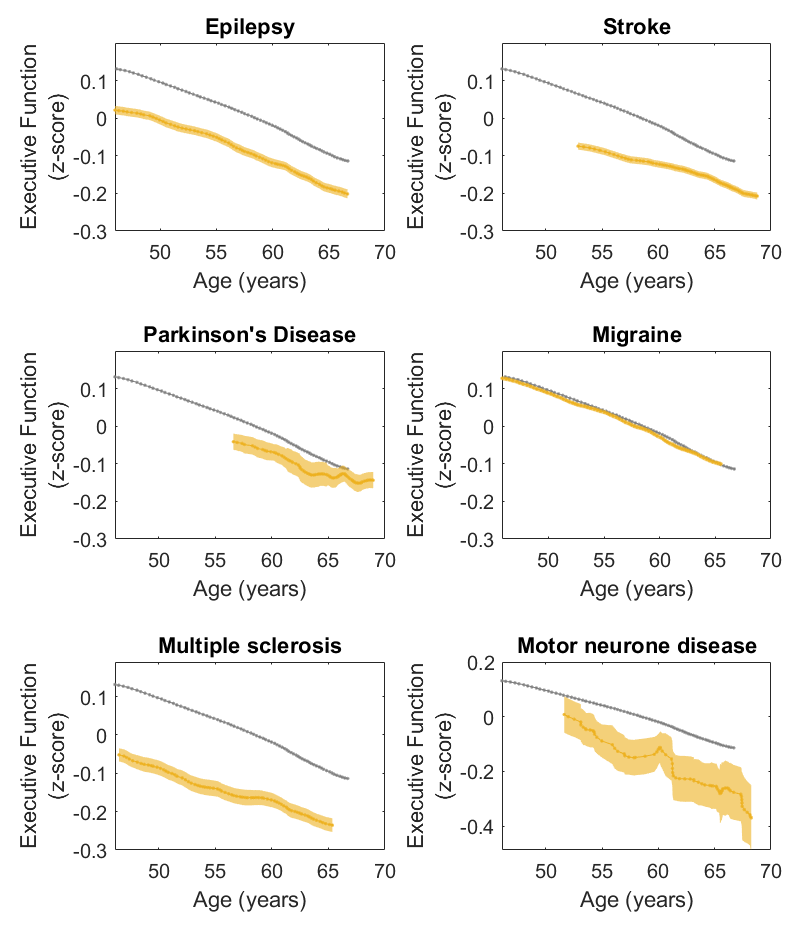

**Figure S4. Executive Function across age for different neurological conditions**

Mean Executive Function declines across age for each neurological condition (yellow) compared with healthy controls (grey). Individuals with epilepsy, stroke, Parkinson’s disease and multiple sclerosis have lower Executive Function for a given age. Error bars denote standard error.

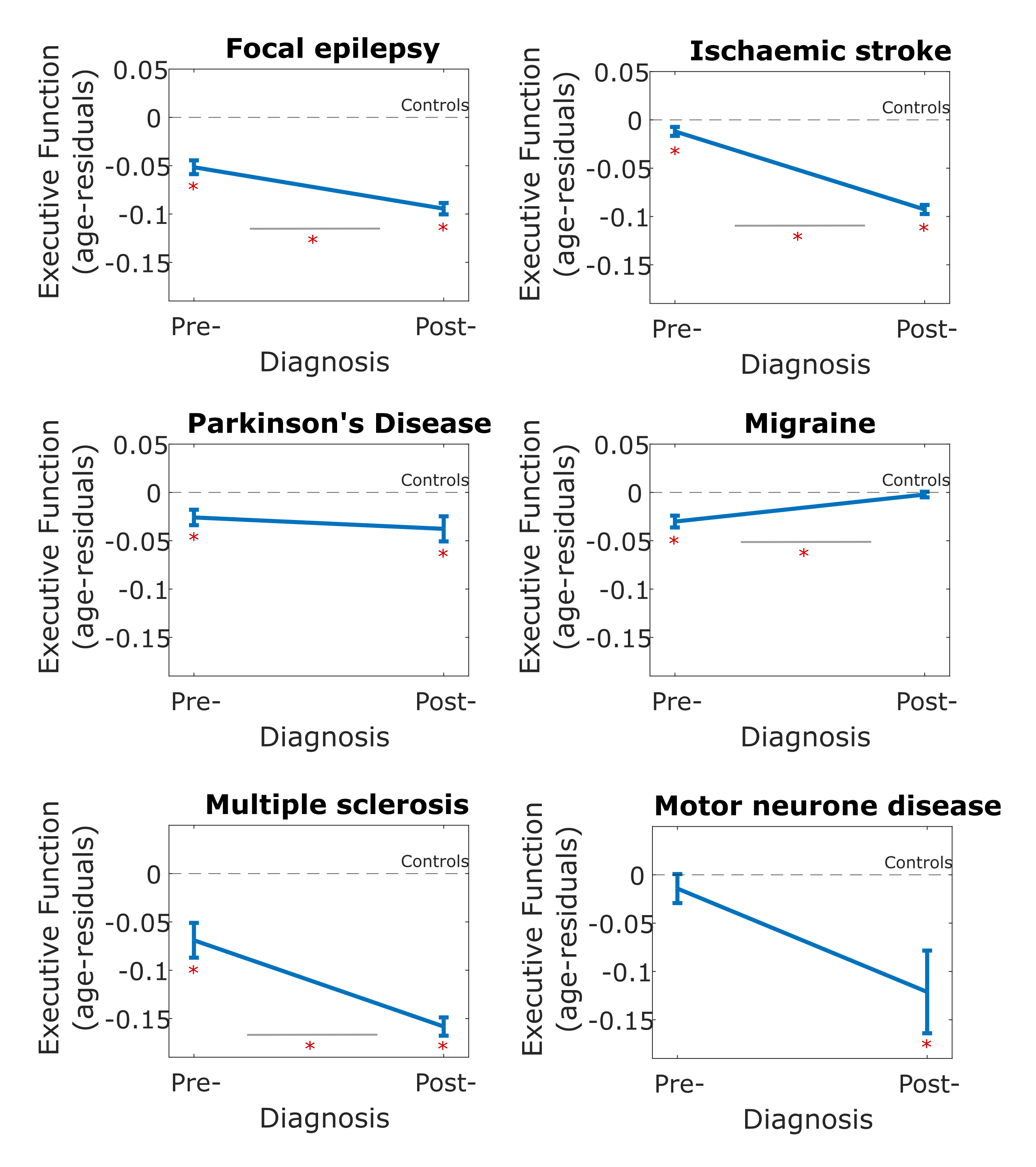

**Figure S5. Comparing pre-diagnosis and post-diagnosis Executive Function for each individual neurological condition**

Executive Function was lower in post-diagnosis participants in all neurological conditions compared to pre-diagnosis participants apart from migraine and motor neurone disease. The level of pre-diagnosis Executive Function was lower than controls in all conditions apart from motor neurone disease. Mean executive function for each condition was compared with control group, asterix (*) below each error bar denotes significant difference p<0.05, while within-condition pre-diagnosis vs. post-diagnosis executive function was compared (represented by an * below grey bars) using a post-hoc Tukey analysis to account for multiple comparison. Error bars denote standard error.

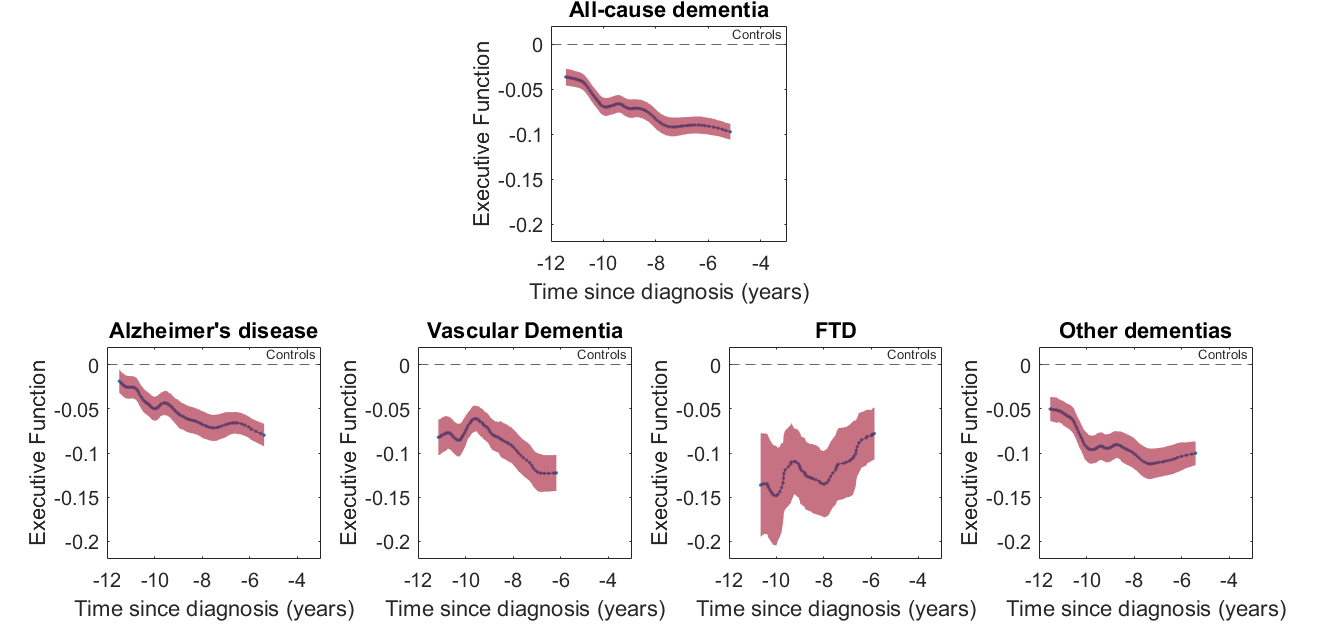

**Figure S6. Pre-diagnosis cognitive profile for participants with dementia**

Executive Function declines progressively leading up to diagnosis of all-cause dementia as well as subtypes of Alzheimer’s disease, vascular dementia and other dementias. Fronto-temporal dementia does not show this change.
